# Contractile and Hemodynamic Modulation of Skeletal Muscle Viscoelasticity Quantified In Vivo by Ultrasound Time-Harmonic Elastography

**DOI:** 10.64898/2026.06.25.26356543

**Authors:** Tom Meyer, Eduard Kurz, Stefan Klemmer Chandía, Pascal Engl, Giacomo Valli, Yanglei Wu, Klaus Jenderka, Thomas Bartels, René Schwesig, Jing Guo, Ingolf Sack, Hossein S. Aghamiry

## Abstract

Skeletal muscle is a living, perfused soft tissue whose viscoelastic behavior is shaped by both voluntary contraction and hemodynamic state. However, the independent and superimposed contributions of contractile loading and blood flow restriction (BFR) have not been quantified simultaneously in real time. Twenty-six healthy adults underwent multi-frequency ultrasound time-harmonic elastography (THE, 60–80 Hz) of the vastus lateralis under six conditions: rest, 15% and 30% maximal voluntary contraction (MVC) before BFR, passive BFR after 4 min of cuff inflation, and 15% and 30% MVC shortly after cuff release. Shear wave speed (SWS), reflecting elasticity, and penetration rate (PR), reflecting inverse viscous damping, were extracted using the k-MDEV inversion algorithm. BFR significantly elevated SWS at all three contraction levels relative to the corresponding pre-BFR measurements (Holm-corrected p ≤ 0.011; dz = 0.54–2.13). PR decreased during resting BFR (dz = 1.34, p < 0.001) and at 15% MVC after cuff release (dz = 0.94, p < 0.001), but not at 30% MVC (dz = 0.21, p = 0.294). BFR-related changes reduced the SWS-force slope by 14.5% and the PR-force slope by 40.7%. Men exhibited a greater BFR-induced increase in resting SWS than women. These findings show that THE can distinguish contractile and hemodynamic contributions to skeletal-muscle viscoelasticity and provide complementary information on elastic and dissipative tissue behavior in vivo.

## 1 Introduction

Quantifying the mechanical state of skeletal muscle under varying physiological conditions is central to skeletal muscle functional assessment. Skeletal muscle is a living, hierarchically organized, and dynamically perfused soft tissue whose mechanical behavior emerges from the interaction of the contractile apparatus, extracellular matrix, and fluid-containing compartments. Shear wave elastography offers a non-invasive way to assess these tissue-level mechanical properties. Elastography exploits the propagation speed and damping of externally induced vibrations or acoustic radiation force [1]. Multiple elastographic modalities have validated shear wave speed (SWS, in m/s) as a reliable surrogate marker of muscle stiffness. Supersonic shear imaging (SSI) [2–4], time-harmonic elastography (THE) [5–7], and magnetic resonance elastography [8] consistently demonstrate that SWS increases with isometric contraction over a broad range of muscle anatomies and contraction intensities. Beyond the elastic response, complex wavenumbers in shear-wave-based elastography provide an attenuation coefficient. Its reciprocal, the penetration rate (PR, in m/s), is related to the distance over which shear waves can penetrate into the tissue. Thus, PR is complementary to SWS, with lower values indicating stronger energy dissipation and greater viscous damping [1, 9, 10]. Simultaneous assessment of SWS and PR may therefore provide a more complete characterization of muscle viscoelasticity than stiffness measurements alone.

While SWS reflects stiffness arising from the contractile apparatus, extracellular matrix, and interstitial and intravascular fluid compartments [1], PR is sensitive to relative motion between the tissue matrix and interstitial or intravascular fluid, and therefore changes with fluid volume and matrix polarity [1, 9]. Voluntary contraction alters both SWS and PR through changes in the contractile apparatus and associated metabolic state. However, the fluid compartments embedded within the tissue can also be modified independently through hemodynamic perturbations, providing a distinct non-contractile contribution to the viscoelastic response.

Blood flow restriction (BFR), achieved by inflating a pneumatic cuff to a fraction of the individual arterial occlusion pressure (AOP), provides a controlled means of modifying the hemodynamic state of the limb without directly altering voluntary motor drive. By occluding venous return while partially preserving arterial inflow, BFR induces venous congestion and blood pooling distal to the cuff and elevates intramuscular pressure (IMP). Increased IMP drives cellular swelling through osmotic shifts and permits accumulation of inorganic phosphate, lactate, and protons [11–13]. These processes are expected to alter the mechanical state of muscle through changes in tissue pressure, fluid distribution, and energy dissipation. However, their collective effects on SWS and PR have not been characterized simultaneously in vivo.

THE is particularly suited to address this question. THE delivers continuous multi-frequency mechanical vibrations externally and records the resulting steady-state wave field [10]. It does not require expensive ultrasound fast scanners and avoids the limitation of acoustic radiation force impulse methods by providing a full field-of-view reconstruction [6, 14, 15]. The k-MDEV inversion [10] utilizes the plane-wave solution of the Helmholtz equation by local phase gradients to estimate SWS and PR from the propagation and attenuation properties of the wave field. This enables complementary elastic and dissipative features of tissue mechanics to be assessed under controlled physiological perturbations.

The present study extends previous work on THE in skeletal muscle by introducing BFR occlusion and post-release conditions, using pre-BFR measurements as within-participant baselines. We investigated how contractile loading and BFR-induced hemodynamic changes jointly affect the viscoelastic behavior of the vastus lateralis (VL) muscle. We tested four hypotheses: (i) passive BFR at 80% AOP elevates VL muscle SWS at rest relative to the pre-BFR condition, indicating non-contractile passive stiffening; (ii) BFR-induced hemodynamic changes elevate SWS above the corresponding pre-BFR value during subsequent isometric contraction, demonstrating superimposition of hemodynamic and contractile stiffness contributions; (iii) both effects are accompanied by reduced PR, reflecting increased viscous damping; and (iv) BFR-induced passive stiffening reduces the SWS-force slope, thereby decreasing the dynamic range of the contractile stiffness response per unit of voluntary force.

## 2 Materials and Methods

### 2.1 Participants

Twenty-six asymptomatic adults (15 men, 11 women) participated after providing written informed consent. Demographic data are summarized in Table 1. All subjects were free of lower-limb musculoskeletal injury, neuromuscular disease, and cardiovascular contraindications to BFR. Participants refrained from vigorous physical activity for 24 h before testing. The study was approved by the ethics committee of Charité – Universitätsmedizin Berlin (EA4/229/25) and by the responsible ethics committee at Martin-Luther-University Halle-Wittenberg (2024-208), and was conducted in accordance with the Declaration of Helsinki.

**Table 1.** Participant demographic characteristics (15 men, 11 women). BMI: body-mass index. Values are reported as mean ± standard deviation and range (Min-Max) for all participants and separately by sex.

| Group | Age (yr) | Height (m) | Mass (kg) | BMI (kg m<sup>-2</sup>) |
| --- | --- | --- | --- | --- |
| Total (n=26) | 25.0±4.1 (20–34) | 1.77±0.09 (1.63–1.92) | 75.5±12.7 (49.8–96.5) | 23.9±2.8 (18.7–30.5) |
| Men (n=15) | 26.8±4.1 (20–34) | 1.83±0.06 (1.69–1.92) | 84.4±7.3 (74.2–96.5) | 25.3±2.3 (20.2–30.5) |
| Women (n=11) | 22.5±2.5 (20–29) | 1.70±0.06 (1.63–1.79) | 63.3±6.8 (49.8–73.0) | 22.0±2.3 (18.7–25.8) |

### 2.2 BFR Protocol and Experimental Design

Participants were seated upright with the hip and knee joints at approximately 90° flexion (Fig. 1). MVC of the right knee extensors was determined in a preliminary session using a load cell (SM-2000N, Interface Inc., Scottsdale, AZ) rigidly attached above the ankle; the higher of two consecutive trials separated by 1 min was retained. AOP was assessed individually by Doppler ultrasound with the cuff positioned at the proximal portion of the right thigh [16]. BFR was applied with the same cuff inflated to 80% of the individually measured AOP. Cuff pressure was monitored continuously by a manometer throughout the BFR condition.

**Fig. 1.**
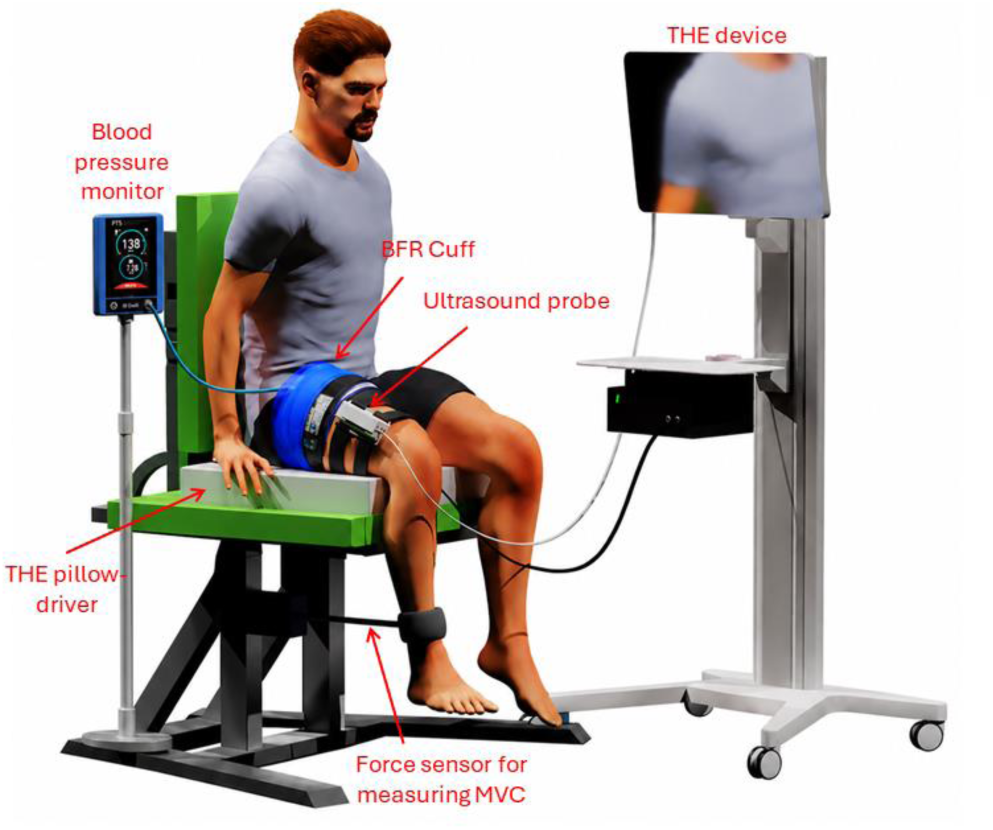
Schematic of the measurement configuration. The participant is seated upright with the right hip and knee at approximately 90°. A custom THE pillow-driver beneath the thigh delivers continuous mechanical vibrations at 60, 70, and 80 Hz. The ultrasound probe is secured over VL at 75% of the distance between the greater trochanter and the lateral femoral epicondyle, aligned parallel to the thigh. A pneumatic cuff at the proximal thigh is inflated to 80% AOP during BFR conditions. A load cell above the ankle provides real-time force feedback.

Six conditions were completed in a fixed sequence. First, all three pre-BFR conditions were obtained with a minimum of 30 seconds rest between measurements while wearing the uninflated cuff:

1. Rest pre-BFR (baseline 12 s THE acquisition),
2. 15% MVC pre-BFR (sustained isometric knee extension at 15% MVC, 12 s THE),
3. 30% MVC pre-BFR (as above, but at 30% MVC).

Following completion of the pre-BFR block, the pneumatic cuff was inflated to 80% AOP. After 4 min of continuous occlusion, a resting BFR measurement was obtained:

4. Rest during passive BFR (12 s THE acquired at the end of the 4-min occlusion).

Approximately 1 min after this acquisition the cuff was deflated. The two post-release conditions were then acquired in immediate succession, without a rest interval between them:

5. 15% MVC post-release (the participant ramped to 15% MVC immediately after cuff deflation), the 12 s THE acquisition began within seconds of release,
6. 30% MVC post-release (within approximately 4 s of completing condition 5), the participant transitioned to 30% MVC for a final 12 s THE acquisition, starting approximately 16 s after cuff deflation.

Real-time knee extension force was displayed to the participant throughout all contraction conditions, allowing participants to follow the requested target force levels (15% and 30% MVC). The full measurement timeline is illustrated in Fig. 2.

**Fig. 2.**
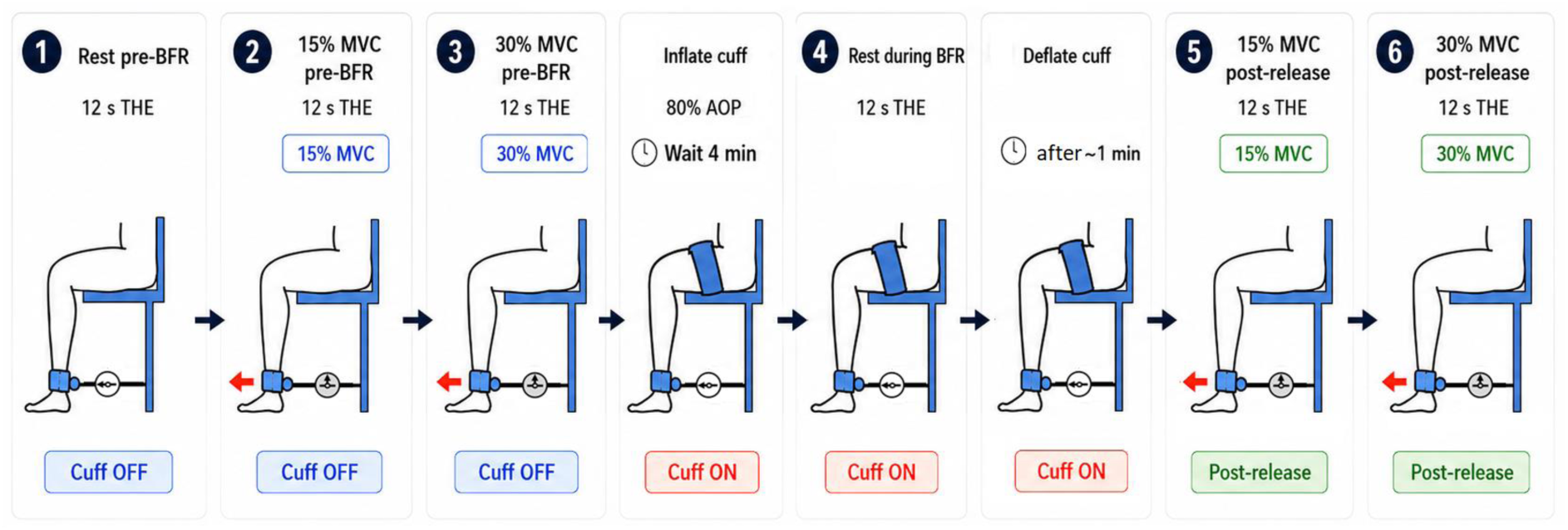
Experimental BFR protocol and timeline. THE measurements were acquired for 12 s at rest and during 15% and 30% MVC before BFR. The cuff was then inflated to 80% AOP, and a resting THE measurement was acquired after 4 min of occlusion. Approximately 1 min later, the cuff was released, followed by THE measurements during 15% and 30% MVC.

### 2.3 Ultrasound Time-Harmonic Elastography

THE was performed as described previously [6, 7]. A custom pillow-driver positioned beneath the participant delivered continuous mechanical vibrations at three superimposed frequencies (60, 70, and 80 Hz). A clinical ultrasound system (GAMPT, Merseburg, Germany) equipped with a linear transducer (LF11-5H60-A3, TeleMed; 5–11 MHz; 54.6 mm footprint) was secured over the VL at 75% of the distance between the greater trochanter and the lateral femoral epicondyle, aligned parallel to the thigh direction (Fig. 1). Radiofrequency (RF) data were acquired line-by-line at 100 frames s⁻¹ for 12 s per condition, yielding 1 200 frames per acquisition.

### 2.4 Data Processing

The k-MDEV inversion algorithm [7, 10] was applied in five sequential steps:

*(1) Displacement estimation:* Axial (vertical) tissue displacements between consecutive RF frames were estimated with a monogenic, phase-constancy optical-flow solver [17, 18]. The RF (or IQ-envelope) data were analyzed in a multiscale band-pass pyramid; at each scale, the monogenic signal (Riesz transform pair plus even component) yielded local amplitude, phase, and orientation in a rotation-invariant representation [17, 18]. Assuming small interframe motion, phase constancy was linearized to relate temporal and spatial monogenic phase gradients to the vertical displacement. The resulting weighted least-squares problem was solved with Tikhonov regularization via Gauss–Newton updates on a coarse-to-fine pyramid [7, Appendix A]. To improve SNR while preserving coherent wave structure, we applied truncated-SVD denoising to the space–time stack, retaining eigenimages of ranks 2–100 [14, 19].
*(2) Frequency realignment:* Aliased frequency components of 60, 70 and 80 Hz at 40, 30, and 20 Hz arising from the 100 Hz frame rate were realigned to the true excitation frequencies using controlled-aliasing reconstruction [10].
*(3) Harmonic isolation:* Third-order Butterworth band-pass filters (±3 Hz bandwidth) isolated each frequency component from the realigned displacement field.
*(4) Directional filtering:* Steerable Gaussian directional filters in the two-dimensional wavenumber domain [20] decomposed each wavefield into eight in-plane propagation directions, suppressing compression artifacts and boundary reflections.
*(5) Wavenumber inversion:* Local wavenumbers were estimated using the k-MDEV framework [10] from the spatial phase gradient. Amplitude-weighted multi-frequency averaging yielded SWS maps at approximately 0.25 mm in-plane resolution. The attenuation coefficient was estimated from the spatial decay of the wave amplitude envelope, and the penetration rate was defined as PR = ω/k″ (m/s), where k″ is the imaginary part of the complex wavenumber. Regions of interest (ROIs) were delineated on the first B-mode frame and vertically shifted in subsequent frames according to vertical correlation with the first frame. Z-score filtering (|z| < 3 on spectral peak sharpness) excluded low-quality pixels [6]. Final SWS and PR values were obtained by temporal and spatial averaging across all 1 200 frames within each ROI.

### 2.5 Statistical Analysis

Normality of paired differences was verified by Shapiro–Wilk tests. Sphericity was assessed by Mauchly’s test with Greenhouse–Geisser correction when violated. All tests were conducted two-sided (α = 0.05). For the manipulation check, a one-way repeated-measures ANOVA tested the effect of contraction level (0%, 15%, 30% MVC) on SWS or PR under pre-BFR conditions, with post-hoc Bonferroni pairwise comparisons.

For the primary contrasts, three pre-planned paired contrasts per outcome variable compared each pre-BFR condition with its BFR or post-release counterpart. Paired t-tests (Wilcoxon signed-rank where normality failed) were applied with Holm–Bonferroni correction within each outcome family (SWS and PR treated separately). Effect sizes are reported as Cohen’s d_z_ with 95% confidence intervals (95% CI) from percentile bootstrap (5 000 resamples).

For the all-vs-rest family, five Holm-corrected paired contrasts tested each non-rest condition against pre-BFR rest.

For the SWS-force and PR-force slope analysis, per-subject linear slopes of SWS and PR on contraction level (0%, 15%, 30% MVC) were computed for the pre-BFR block and the BFR/post-release block separately, then compared by paired t-test. For the sex-stratified slope analysis, within-sex slopes were compared by paired t-test. Between-sex differences in slope values at each block were assessed by Welch’s independent-samples t-test.

For the sex-stratified analysis, BFR-induced deltas (Δ = BFR/post-release − pre-BFR) were compared between the 15 male and 11 female participants by independent-samples t-tests with Benjamini–Hochberg (BH) false discovery rate (FDR) correction within each outcome family (three conditions per outcome). Within-sex effects were assessed by one-sample t-tests on deltas with Holm–Bonferroni correction.

For the correlation analysis, Pearson correlations among age, BMI, SWS, and PR were assessed with BH FDR control (q = 0.05).

Receiver operating characteristic (ROC) curves were constructed separately for each contraction level (rest, 15% MVC, and 30% MVC) to characterize the discriminative capacity of SWS and PR for identifying BFR hemodynamic state by classifying observations as pre-BFR (class 0) or BFR/post-release (class 1). The area under the curve (AUC) was direction-adjusted such that AUC ≥ 0.5 in all cases. Scores were inverted when lower values indicated the BFR/post-release state. AUC confidence intervals (95%) were estimated by subject-level paired bootstrap resampling (5 000 resamples). Optimal classification thresholds were identified by the Youden index. This analysis is presented as an exploratory characterization of state discrimination and does not constitute diagnostic validation.

## 3 Results

Measurements were completed for all 26 participants across all six conditions. Representative SWS and PR maps for a single participant are shown in Fig. 3, illustrating the spatial pattern of BFR- and MVC-induced changes across all six conditions. For this representative participant, resting BFR increased from 2.02 ± 0.43 m/s to 2.96 ± 0.57 m/s, while the corresponding PR decreased from 2.27 ± 0.66 to 2.01 ± 0.71 m/s. At 15% MVC, SWS increased from 3.00 ± 0.43 m/s to 3.75 ± 0.53 m/s post-release, accompanied by a reduction in PR from 1.95 ± 0.60 m/s to 1.25 ± 0.42 m/s. At 30% MVC, SWS increased from 3.86 ± 0.56 m/s to 4.29 ± 0.50 m/s, whereas PR changed only slightly from 1.51 ± 0.46 m/s to 1.27 ± 0.22 m/s.

**Fig. 3.**
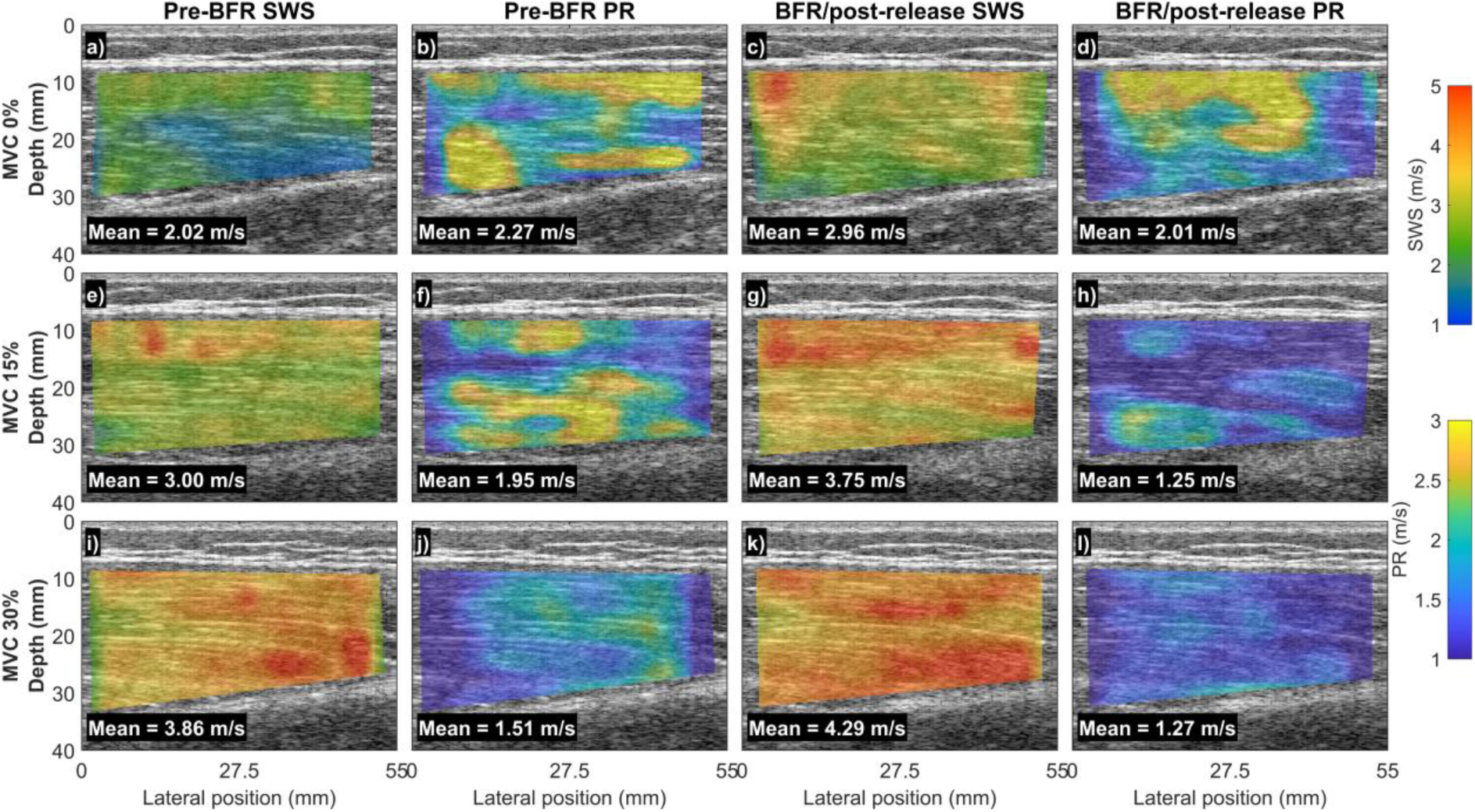
Representative SWS and PR maps overlaid on B-mode images. The maps are shown as semi-transparent overlays on B-mode images within selected regions of interest in the VL muscle across the experimental conditions. Maps are presented for the three contraction levels: rest in the first row, 15% MVC in the second row, and 30% MVC in the third row. Pre-BFR conditions are shown in columns 1 and 2, whereas BFR/post-release conditions are shown in columns 3 and 4. SWS maps are shown in columns 1 and 3, and PR maps are shown in columns 2 and 4. The maps are visualized only within the ROI, while the underlying B-mode image provides anatomical reference. Color bars are kept identical across comparable maps to facilitate visual comparison across conditions.

Group SWS increased monotonically from 2.45 ± 0.15 m/s at rest pre-BFR to 3.90 ± 0.35 m/s at 30% MVC post-release, reflecting the combined contributions of voluntary contraction and BFR-induced hemodynamic changes. Over the same progression, PR declined from 1.87 ± 0.30 m/s at rest pre-BFR to 1.30 ± 0.20 m/s at 30% MVC post-release, with a transient reversal between resting BFR and 15% MVC pre-BFR. One reason for such instability may be the lower accuracy of the estimated PR compared to SWS. Full descriptive statistics are given in Table 2, while individual parameter changes are shown in Fig. 4a-b. In addition, changes relative to pre-BFR rest are shown in Fig. 5.

**Fig. 4a-b.**
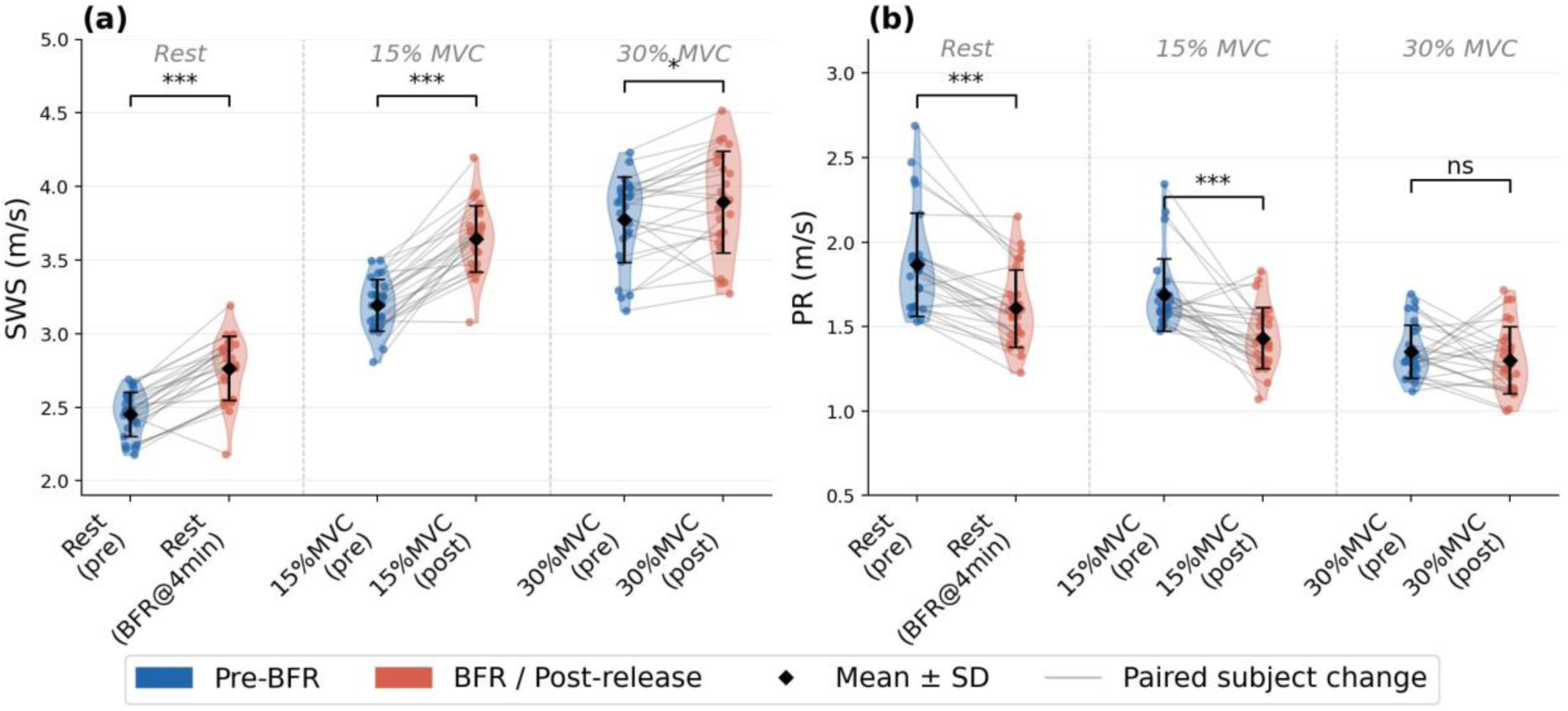
VL biomechanical properties across MVC and BFR phases. Panel (a) shows SWS and panel (b) shows PR. Violin plots indicate the distribution of individual measurements across all six conditions. Blue distributions represent pre-BFR measurements, while the red distributions represent during BFR or post-release measurements. Individual data points are shown with circles and filled diamonds, black vertical lines indicate group mean ± standard deviation, and gray lines connect paired observations within the same participant. Significance brackets indicate Holm-corrected paired contrasts between corresponding pre-BFR and BFR/post-release conditions (***p < 0.001, *p < 0.05).

**Fig. 5a-b.**
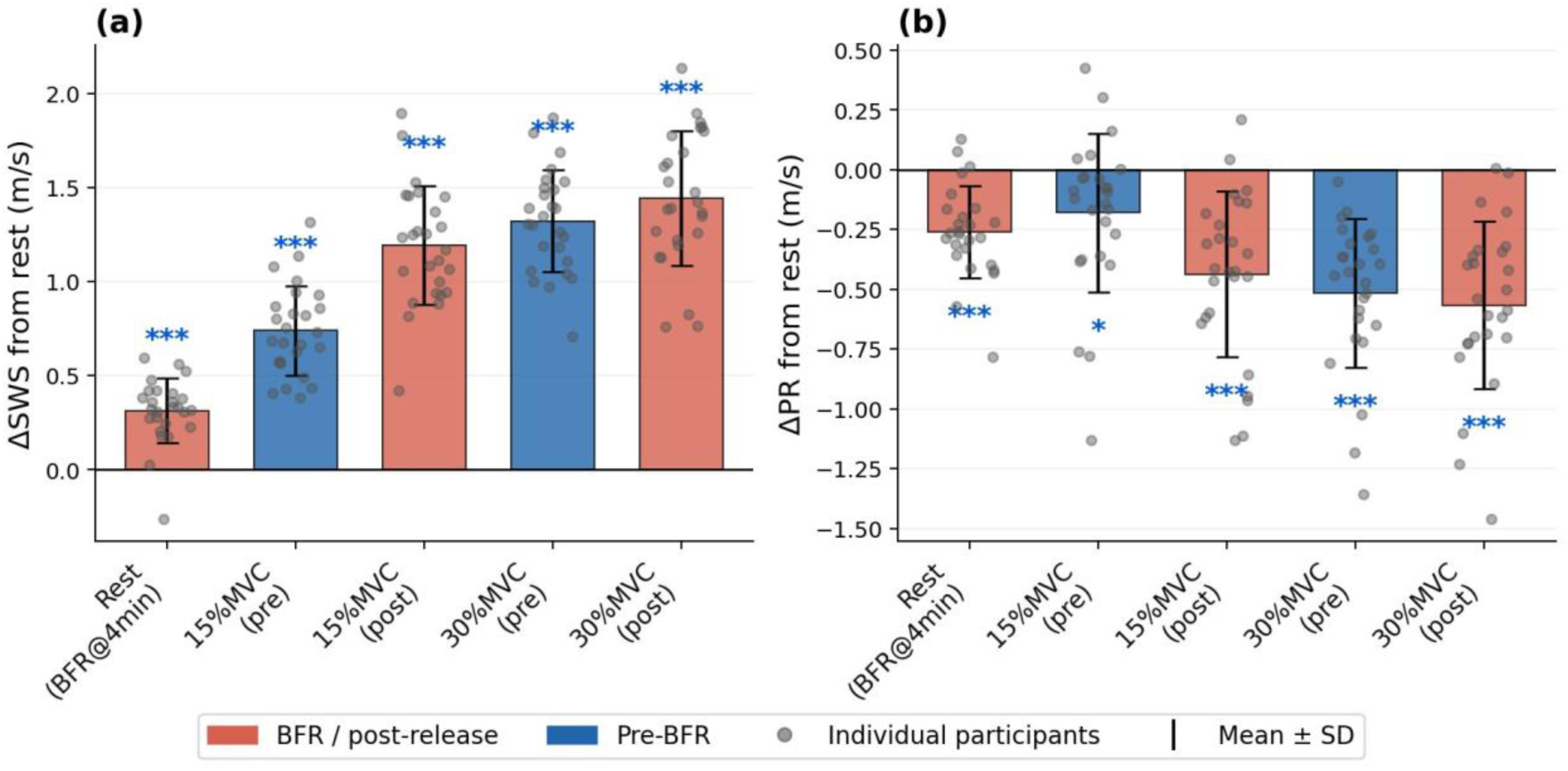
Change in VL biomechanical parameters relative to pre-BFR rest. Panel (a) shows the change in SWS and panel (b) shows the change in PR for the five remaining conditions. Bars indicate group mean change from pre-BFR rest, vertical error bars indicate ± standard deviation, and gray circles show individual participant values. Red bars represent during BFR or post-release conditions, while blue bars represent pre-BFR contraction conditions. Significance asterisks denote Holm-corrected paired comparisons versus pre-BFR rest (***p < 0.001, *p < 0.05).

**Table 2.** Descriptive statistics for SWS and PR across six conditions. Values are mean ± SD [min-max].

| Condition |  | SWS (m/s) | PR (m/s) |
| --- | --- | --- | --- |
| Rest | Pre-BFR | $2.45 \pm 0.15$ [2.18, 2.69] | $1.87 \pm 0.31$ [1.53, 2.69] |
| | BFR @ 4 min | $2.77 \pm 0.22$ [2.18, 3.19] | $1.61 \pm 0.23$ [1.23, 2.15] |
| 15% MVC | Pre-BFR | $3.19 \pm 0.18$ [2.81, 3.50] | $1.69 \pm 0.21$ [1.47, 2.34] |
| | Post-release | $3.65 \pm 0.23$ [3.08, 4.20] | $1.43 \pm 0.18$ [1.07, 1.83] |
| 30% MVC | Pre-BFR | $3.78 \pm 0.29$ [3.15, 4.23] | $1.35 \pm 0.16$ [1.12, 1.69] |
| | Post-release | $3.90 \pm 0.35$ [3.27, 4.52] | $1.30 \pm 0.20$ [1.00, 1.71] |

### 3.1 Pre-BFR Results

Before testing BFR effects, we verified that the contraction task produced the expected dose-dependent stiffness response under normal perfusion.

A one-way repeated-measures ANOVA showed a strong effect of contraction level on SWS. This effect was significant (*F*(2, 50) = 311.91, *p* < 0.001, η_p_ ^2^ = 0.926; sphericity upheld, *W* = 0.935, *p* = 0.446).

Post-hoc comparisons confirmed SWS increases at each step. SWS increased from rest to 15% MVC (Δ = +0.74 m/s, *g* = 4.46, *p* < 0.001) and from 15% to 30% MVC (Δ = +0.58 m/s, *g* = 2.39, *p* < 0.001).

This pattern is consistent with prior SSI studies of the VL at comparable contraction levels [3, 4, 21]. For PR, the ANOVA was also significant (*F*(2, 50) = 45.16, *p* < 0.001, η_p_ ^2^ = 0.644; sphericity upheld, *W* = 0.970, *p* = 0.698). PR decreased significantly from rest to both contraction levels. This included rest vs 15% MVC (*g* = 0.55, *p* = 0.010) and rest vs 30% MVC (*g* = 1.66, *p* < 0.001). PR also showed a significant further decrease from 15% to 30% MVC (*g* = 1.18, *p* < 0.001).

The progressive decline in PR across force levels contrasts with the continuously rising SWS. This demonstrates that the elastic and viscous components carry non-redundant mechanical information. It also shows that they exhibit distinct force-level dependencies, a finding examined further in the Discussion.

### 3.2 BFR-induced Changes

BFR significantly elevated SWS in all three condition pairs after Holm–Bonferroni correction (Table 3), verifying hypotheses (i) and (ii). The largest increase occurred at 15% MVC (+0.45 ± 0.21 m/s; *d*_z_ = 2.13, 95% CI [1.59, 3.17]), an intermediate effect occurred at rest (+0.31 ± 0.17 m/s; *d*_z_ = 1.83 [1.09, 3.57]), whereas the smallest increase occurred at 30% MVC (+0.12 ± 0.22 m/s; *d*_z_ = 0.54 [0.13, 1.27], *p*(Holm) = 0.011). This pattern may also reflect the timing of the acquisitions. The 15% MVC acquisition began within seconds of cuff deflation, whereas the 30% MVC acquisition started approximately 20 s later, potentially during ongoing hemodynamic recovery.

**Table 3.** Primary BFR-induced paired contrasts, Holm–Bonferroni corrected within each outcome family. Δ is defined as BFR/post-release minus pre-BFR (positive = increase). dz: Cohen’s dz with 95% bootstrap CI (5 000 resamples).

| Parameter | Condition | $\Delta$ (mean ± SD) | t | p(Holm) | $d_z$ [95% CI] |
| --- | --- | --- | --- | --- | --- |
| SWS (m/s) | Rest: pre vs. BFR @ 4 min | +0.31 ± 0.17 | –9.33 | <0.001 *** | 1.83 [1.09, 3.57] |
| | 15% MVC: pre vs. post-release | +0.45 ± 0.21 | –10.88 | <0.001 *** | 2.13 [1.59, 3.17] |
| | 30% MVC: pre vs. post-release | +0.12 ± 0.22 | –2.75 | 0.011 * | 0.54 [0.13, 1.27] |
| PR (m/s) | Rest: pre vs. BFR @ 4 min | –0.26 ± 0.19 | +6.83 | <0.001 *** | 1.34 [0.93, 2.16] |
| | 15% MVC: pre vs. post-release | –0.26 ± 0.27 | +4.81 | <0.001 *** | 0.94 [0.61, 1.44] |
| | 30% MVC: pre vs. post-release | –0.05 ± 0.24 | +1.07 | 0.294 (ns) | 0.21 [–0.17, 0.68] |

PR decreased significantly at rest (*d*_z_ = 1.34, *p*(Holm) < 0.001) and at 15% MVC post-release (*d*_z_ = 0.94, *p*(Holm) < 0.001, Table 3). This partially verifies hypothesis (iii). The 30% MVC contrast did not reach significance (*d*_z_ = 0.21, *p*(Holm) = 0.294).

Significant PR reductions were therefore observed at rest. At this level, no contractile activity was present. Significant reductions were also observed at 15% MVC post-release. This acquisition was captured within seconds of cuff deflation at the onset of reactive hyperemia. PR did not decrease significantly at 30% MVC. This occurred despite a substantially lower absolute baseline (1.35 m/s vs. 1.69 m/s at 15% MVC pre-BFR). This finding is consistent with the reduced dynamic range at high contraction intensity. It is also consistent with the approximately 20 s elapsed since cuff deflation by the time of that acquisition. During this interval, partial hemodynamic recovery would already be expected.

The non-parallel behavior of SWS and PR across conditions suggests that both parameters reflect mechanistically distinct tissue properties, depending on force level and hemodynamic state.

### 3.3 Effect Sizes Relative to Pre-BFR Rest

All five non-rest conditions differed significantly from pre-BFR rest (Holm-corrected, *p* < 0.001 for all ten contrasts). Effect sizes are reported in Table 4. For SWS, *d*_z_ increased progressively from 1.83 (BFR at rest) to 4.85 (30% MVC pre-BFR), reflecting the additive contributions of passive hemodynamic stiffening and voluntary contraction. For PR, *d*_z_ ranged from 0.55 (15% MVC pre-BFR) to 1.66 (30% MVC pre-BFR), with the BFR-state conditions showing intermediate values (1.34 at resting BFR; 1.26 at 15% MVC post-release).

**Table 4.**
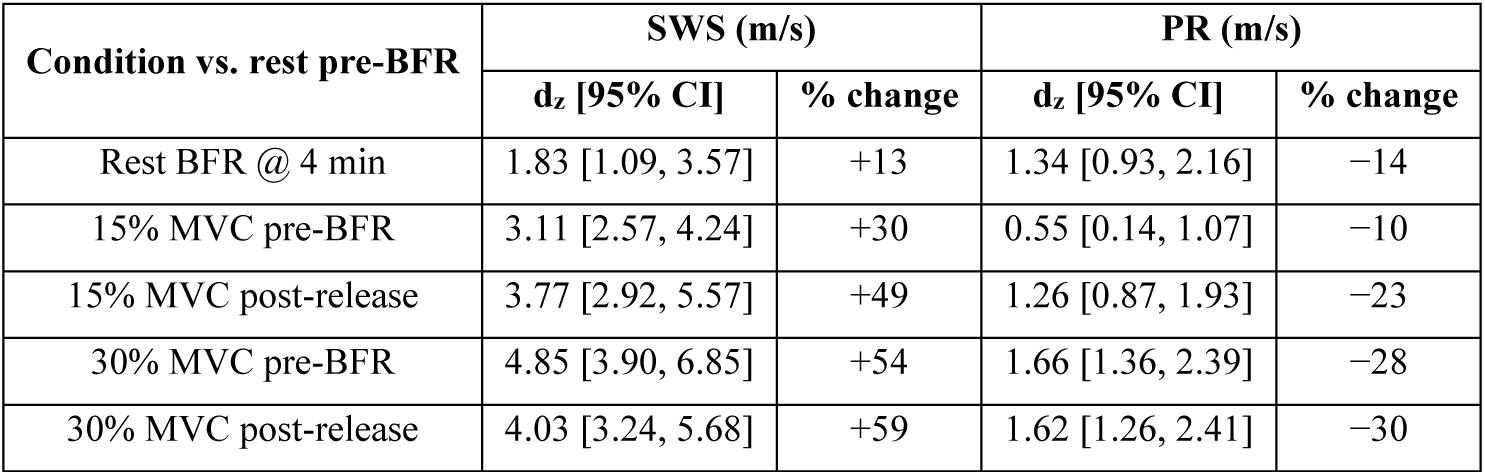
Effect sizes (Cohen’s dz) for all five conditions versus pre-BFR rest (all Holm-corrected p < 0.001). For SWS, positive dz indicates an increase; for PR, positive dz indicates a decrease.

| Condition vs. rest pre-BFR | SWS (m/s) |  | PR (m/s) |  |
| --- | --- | --- | --- | --- |
| | $d_z$ [95% CI] | % change | $d_z$ [95% CI] | % change |
| Rest BFR @ 4 min | 1.83 [1.09, 3.57] | +13 | 1.34 [0.93, 2.16] | –14 |
| 15% MVC pre-BFR | 3.11 [2.57, 4.24] | +30 | 0.55 [0.14, 1.07] | –10 |
| 15% MVC post-release | 3.77 [2.92, 5.57] | +49 | 1.26 [0.87, 1.93] | –23 |
| 30% MVC pre-BFR | 4.85 [3.90, 6.85] | +54 | 1.66 [1.36, 2.39] | –28 |
| 30% MVC post-release | 4.03 [3.24, 5.68] | +59 | 1.62 [1.26, 2.41] | –30 |

### 3.4 Force–Stiffness Slope Analysis

Per-subject SWS-force slopes were significantly steeper in the pre-BFR block (0.04 ± 0.01 m/s per % MVC) than in the BFR/post-release block (0.04 ± 0.01 m/s per % MVC; paired *t*-test: *t*(25) = 3.46, *p* = 0.002, *d*_z_ = 0.68 [0.26, 1.13]; Fig. 6a). This 15% slope reduction confirms hypothesis (iv), indicating that BFR-induced passive stiffening increases SWS and reduces the dynamic range available for the contractile increment per unit voluntary force.

**Fig. 6a-b.**
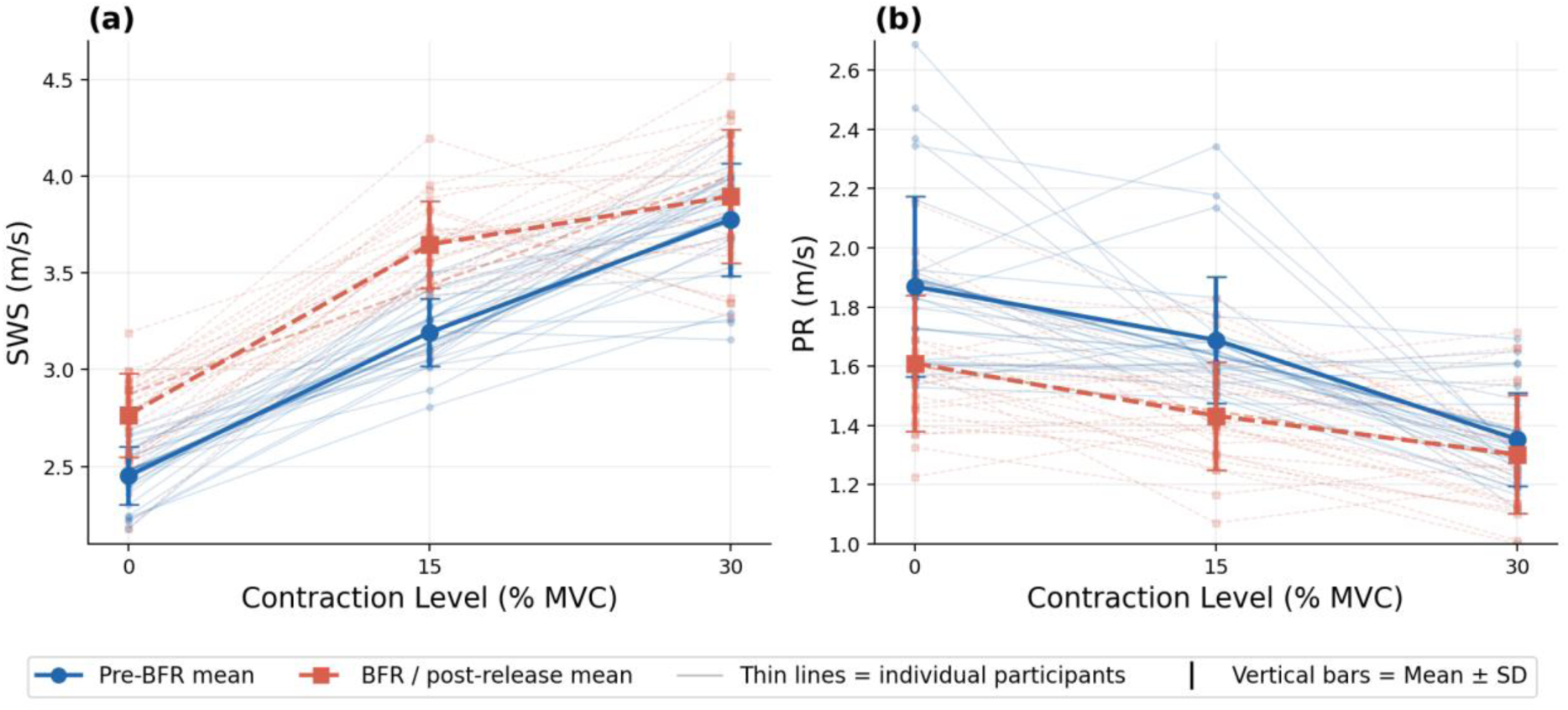
Force–viscoelasticity relationships before and after blood flow restriction. Panel (a) shows SWS and panel (b) shows PR, each plotted as a function of contraction level (0%, 15%, and 30% MVC). Thin semi-transparent lines represent individual participant trajectories, thick lines with markers indicate group means with ± standard deviation error bars. Blue solid lines represent pre-BFR measurements and red dashed lines represent BFR/post-release measurements.

The PR-force slope was similarly attenuated under BFR. Pre-BFR, PR declined with increasing contraction level at a mean rate of −0.02 ± 0.01 m/s per % MVC; post-BFR, this rate was reduced in magnitude to −0.01 ± 0.01 m/s per % MVC (paired *t*-test: *t*(25) = 3.36, *p* = 0.003, *d*_z_ = 0.66 [0.31, 1.12]; Fig. 6b), representing a 41% reduction in absolute slope magnitude. As with SWS, BFR-induced PR suppression at rest shifted the hemodynamic baseline and reduced the dynamic range available for the contraction-dependent PR response.

### 3.5 Correlations

SWS and PR were not significantly correlated at rest, at 15% MVC pre-BFR, at 15% MVC post-release, or during resting BFR (*r* = −0.04 to +0.03, all *p* > 0.85). Marginal negative associations appeared at 15% MVC pre-BFR (*r* = −0.355, *p* = 0.075) and at 30% MVC post-release (*r* = −0.402, *p* = 0.042), but neither survived BH correction. No FDR-significant SWS-PR correlation was found at any condition. Neither BMI nor age correlated significantly with SWS after FDR correction (*r* = 0.027 and *r* = 0.217, respectively; both *p* > 0.08).

### 3.6 Sex-Stratified Analysis

BFR-induced deltas were compared between the 15 male and 11 female participants. Full results are given in Table 5 and Fig. 7.

**Fig. 7a-d.**
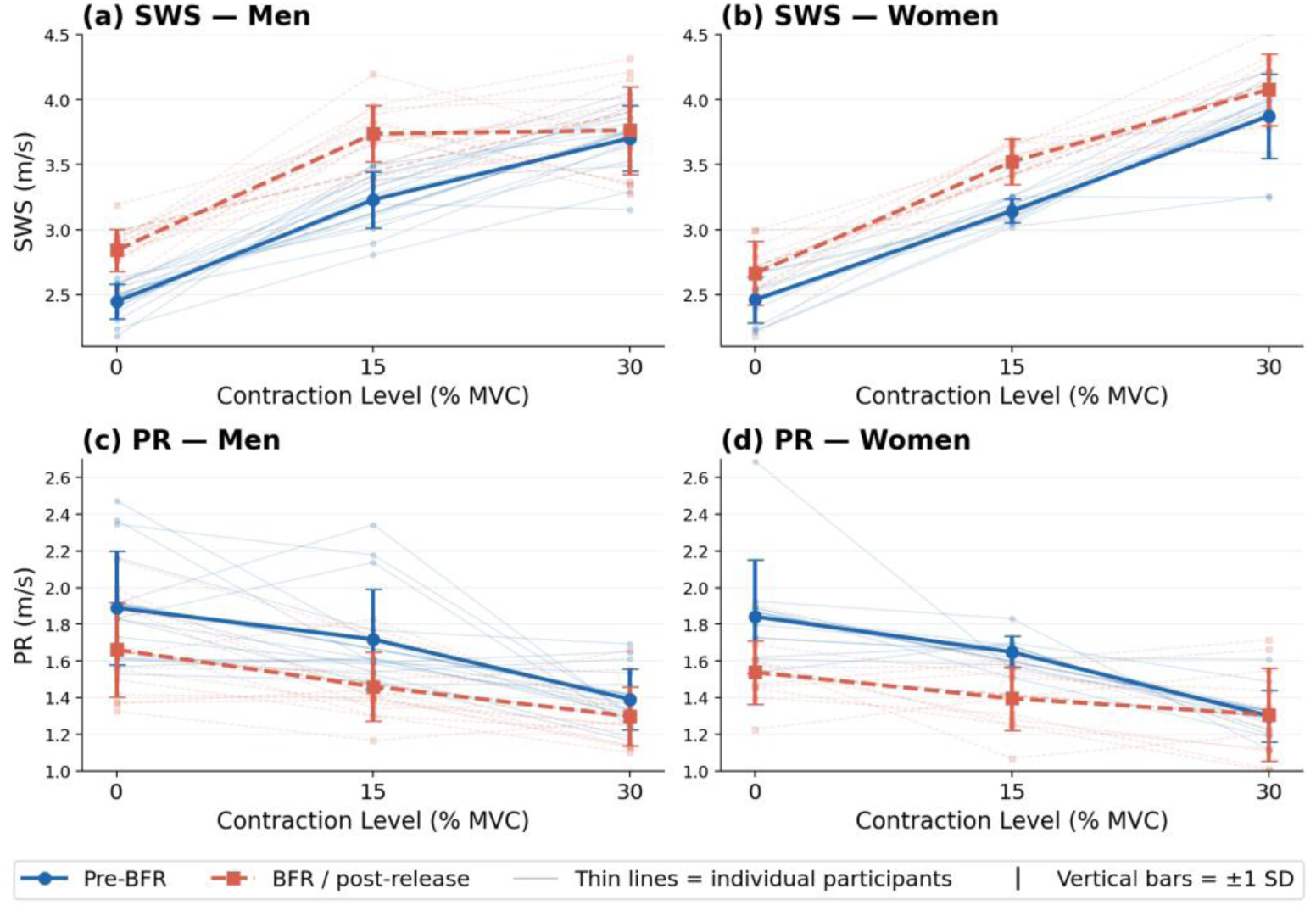
Sex-stratified biomechanical response to blood flow restriction. Panels (a) and (b) show SWS in men (N = 15) and women (N = 11), respectively. Panels (c) and (d) show PR in men and women, respectively. Values are plotted across 0%, 15%, and 30% MVC. Blue solid lines indicate pre-BFR measurements and red dashed lines indicate BFR/post-release measurements. Thin semi-transparent lines represent individual participant trajectories, and thick lines with markers indicate group means with ± standard deviation error bars.

**Table 5.** Sex-stratified BFR-induced changes in SWS and PR (men N = 15, women N = 11). Δ: BFR/post-release minus pre-BFR (mean ± SD). dz: within-sex Cohen’s dz (95% bootstrap CI). p(within): Holm–Bonferroni corrected one-sample t-test. p(sex): BH-FDR-corrected between-sex test; d: Cohen’s d. ***p < 0.001, **p < 0.01, *p < 0.05.

| Parameter | Condition | Sex | $\Delta$ (mean ± SD) | dz [95% CI] | $p(\text{within})$ | $p(\text{sex})$ [d] |
| --- | --- | --- | --- | --- | --- | --- |
| SWS (m/s) | Rest | Men | +0.40±0.11 | 3.48 [2.70, 5.58] | <0.001 *** | 0.008 ** [1.32] |
| | | Women | +0.20±0.18 | 1.13 [0.36, 5.43] | 0.008 ** | |
| | 15% MVC | Men | +0.51±0.23 | 2.18 [1.57, 3.51] | <0.001 *** | 0.133 [0.62] |
| | | Women | +0.38±0.16 | 2.32 [1.35, 11.80] | <0.001 *** | |
| | 30% MVC | Men | +0.06±0.24 | 0.25 [−0.25, 1.23] | 0.352 | 0.102 [−0.67] |
| | | Women | +0.20±0.18 | 1.13 [0.58, 2.80] | 0.008 ** | |
| PR (m/s) | Rest | Men | −0.23±0.18 | −1.24 [−2.52, −0.73] | <0.001 *** | 0.337 [0.39] |
| | | Women | −0.30±0.21 | −1.45 [−4.29, −0.91] | 0.002 ** | |
| | 15% MVC | Men | −0.26±0.31 | −0.84 [−1.53, −0.41] | 0.012 * | 0.970 [−0.01] |
| | | Women | −0.25±0.23 | −1.11 [−1.93, −0.77] | 0.008 ** | |
| | 30% MVC | Men | −0.09±0.24 | −0.38 [−1.10, 0.12] | 0.161 | 0.320 [−0.40] |
| | | Women | +0.01±0.24 | 0.02 [−0.89, 0.60] | 0.942 | |

At rest, men exhibited a BFR-induced SWS elevation of +0.40 ± 0.11 m/s, nearly double that of women (+0.20 ± 0.18 m/s; *t*(24) = 3.34, *q*_BH_ = 0.008, *d* = 1.32). This was the only between-sex contrast for ΔSWS that survived FDR correction; differences at 15% MVC (*d* = 0.62, *q* = 0.133) and 30% MVC (*d* = 0.67, *q* = 0.132) were not significant. Within-sex patterns differed qualitatively across force levels: men showed large, significant BFR-induced SWS elevations at rest (*d*_z_ = 3.48, *p*(Holm) < 0.001) and at 15% MVC (*d*_z_ = 2.18, *p*(Holm) < 0.001) but not at 30% MVC (*d*_z_ = 0.25, *p*(Holm) = 0.352), whereas women showed significant SWS elevations at all three force levels (*d*_z_ = 1.13–2.32; all *p* ≤ 0.008).

Within-sex PR effects were broadly similar in direction and magnitude between men and women. No between-sex contrast in ΔPR survived FDR correction (all *q*_BH_ ≥ 0.32, all *d* ≤ 0.40).

In men, the BFR/post-release slope (0.03 ± 0.01 m/s per % MVC) was 27% shallower than the pre-BFR slope (0.04 ± 0.01 m/s per % MVC; *t*(14) = 5.78, *p* < 0.001, *d*_z_ = 1.49 [1.19, 2.19]), while in women the two slopes were identical (0.047 ± 0.008 m/s vs. 0.047 ± 0.010 m/s per % MVC; *t*(10) = −0.01, *p* = 0.993, *d*_z_ = 0.00 [−0.54, 1.02]). Accordingly, the sex difference in SWS slope values under BFR was significant (*t* = −3.64, *p* = 0.001; Fig. 7a-d).

In women, the BFR/post-release PR slope (−0.01 ± 0.01 m/s per % MVC) was significantly shallower than the pre-BFR slope (−0.02 ± 0.01 m/s per % MVC; *t*(10) = 3.76, *p* = 0.004, *d*_z_ = 1.13). In men, the corresponding change was not significant (−0.01 ± 0.01 m/s vs. −0.02 ± 0.01 m/s per % MVC; *t*(14) = 1.56, *p* = 0.141, *d*_z_ = 0.40). The sex difference in PR slope values at BFR did not reach significance (*t* = −1.13, *p* = 0.274).

### 3.7 Exploratory Receiver Operating Characteristic (ROC) Analysis

ROC analysis examined if SWS and PR can discriminate pre-BFR from BFR/post-release state at each contraction level (Fig. 8). For SWS, discrimination was excellent at 15% MVC (AUC = 0.95, 95% CI [0.87, 0.99]) and strong at rest (AUC = 0.88 [0.79, 0.95]), but weak at 30% MVC (AUC = 0.61 [0.52, 0.70]). For PR, which decreases under BFR state, direction-adjusted AUC was strong at 15% MVC (AUC = 0.85 [0.74, 0.95]) and moderate at rest (AUC = 0.76 [0.69, 0.86]), with poor discrimination at 30% MVC (AUC = 0.59 [0.45, 0.73]). The progressive degradation of AUC from 15% MVC to 30% MVC for both parameters is consistent with the reduction of BFR-induced changes at high contraction intensities observed in the primary contrasts. Youden-optimal thresholds for discriminating BFR/post-release from pre-BFR using SWS were ≥ 2.70 m/s (rest; sensitivity 0.69 [95% CI: 0.50, 0.85], specificity 1.00 [1.00, 1.00]) and ≥ 3.40 m/s (15% MVC; sensitivity 0.92 [95% CI: 0.81, 1.00], specificity 0.85 [0.69, 0.96]).

**Fig. 8a-b.**
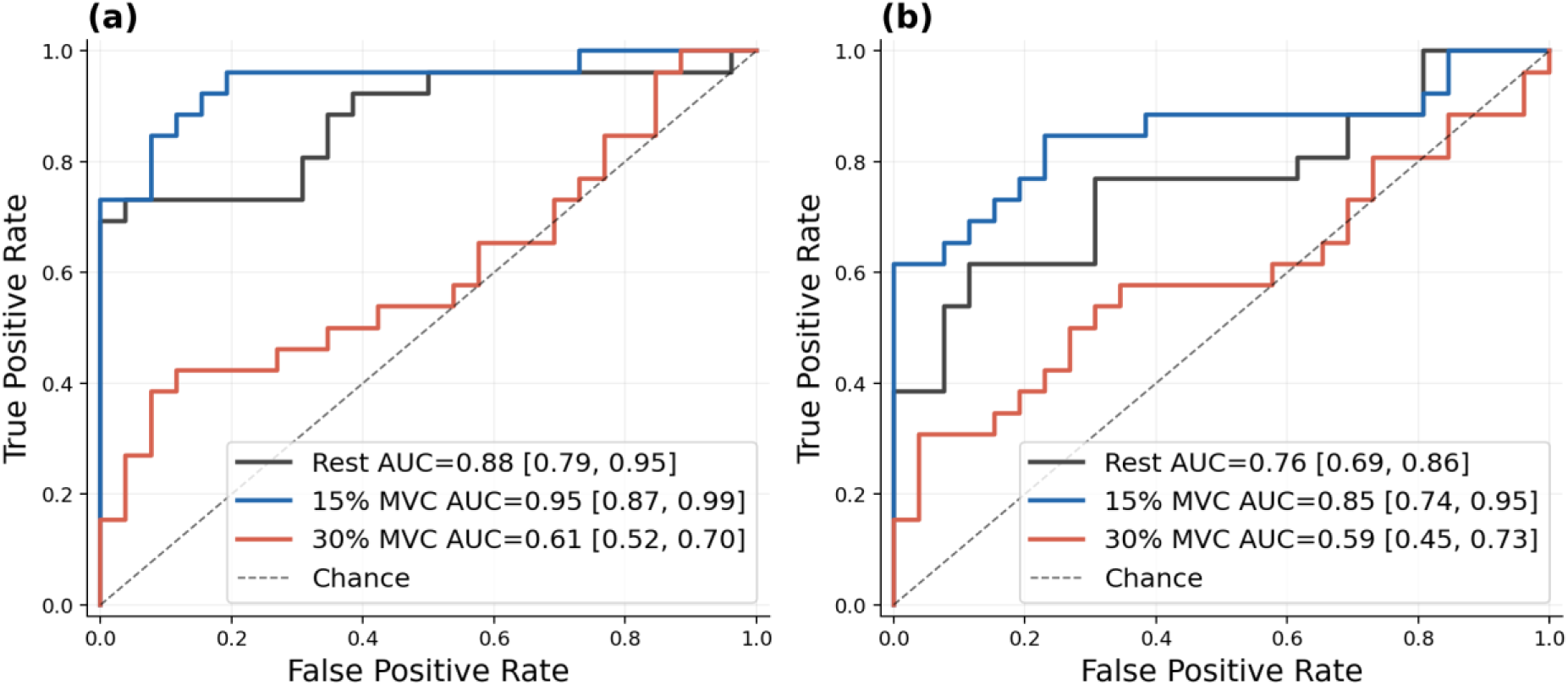
ROC analysis for discriminating pre-BFR from BFR/post-release state. Panel (a) shows ROC curves for SWS and panel (b) shows ROC curves for PR. Separate curves are shown for rest (black), 15% MVC (blue), and 30% MVC (red). AUC values are direction-adjusted so that values above 0.5 indicate discrimination between experimental states; 95% confidence intervals were estimated by subject-level paired bootstrap resampling (5 000 resamples). The diagonal dashed line represents chance-level discrimination.

## 4 Discussion

To the best of our knowledge, this is the first study to investigate VL viscoelasticity at rest and during active muscle contraction, both with and without passive BFR, while accounting for sex-specific effects. The results highlight two main points. First, under normal perfusion, SWS increased stepwise with contraction, whereas PR decreased, indicating that muscle activation increases the elastic component while also increasing viscous damping. This is consistent with prior muscle elastography studies showing a near-linear rise in shear stiffness with isometric load [7, 21, 22]. Second, hemodynamic obstruction altered muscle viscoelasticity even in the absence of voluntary contraction. Together, these findings indicate that the mechanical behavior of skeletal muscle reflects not only active force generation but also the state of its fluid-containing compartments.

The activation-dependent increase in SWS can be interpreted within the framework proposed by Schrank et al. [23], in which muscle contraction changes the molecular and structural interactions within the contractile apparatus. Calcium-mediated actin-myosin cross-bridging, filament sliding, and the associated reduction in local hydration and lubrication zones may increase mechanical coupling and intra-fibrous friction (Fig. 7 of [23]). In the present study, this mechanism is reflected by the monotonic increase in SWS and the decrease in PR during the muscle states without BFR. These responses are consistent with the view that active contraction modifies both the elastic and dissipative behavior of the tissue.

During resting BFR, SWS increased by approximately 13% and PR decreased by 14% relative to pre-BFR rest. BFR likely adds a different, non-contractile mechanism: venous congestion, vessel-wall expansion, and increased intramuscular fluid pressure may pre-stress the muscle as a fluid-filled composite tissue, producing strain stiffening even without active force generation. Changes in the distribution and pressure of the intravascular and interstitial fluid compartments may also alter energy dissipation through fluid–matrix interactions. This pressure-related stiffening is consistent with elastography observations in other organs, where increased vascular or compartmental pressure is accompanied by higher tissue stiffness, including recent work linking liver stiffness and volumetric strain to portal pressure [24–26].

The post-release contraction measurements support this interpretation. Relative to pre-BFR rest, SWS increased by 30% during 15% MVC before BFR and by 48.7% during 15% MVC immediately after cuff release. PR decreased by 10% and 23%, respectively. Because resting BFR alone changed SWS and PR by 13% and 14% in the absence of voluntary contraction, the larger post-release response at 15% MVC is unlikely to reflect greater contractile activation alone compared with pre-BFR at 15% MVC. Instead, it supports a superimposed hemodynamic contribution that adds to the activation-dependent change. This interpretation is also consistent with reports that muscle mechanical properties change rapidly under BFR and recover after cuff release [27].

SWS remained higher than the corresponding pre-BFR values at both post-release contraction levels, but the BFR-related increase was larger at 15% MVC than at 30% MVC (+0.45 m/s vs. +0.12 m/s). This difference may reflect the timing of acquisition: the 15% MVC measurement started within seconds of cuff release, whereas the 30% MVC measurement followed roughly 20 s later. The smaller effect at 30% MVC may therefore reflect partial hemodynamic recovery during early reperfusion [27], together with a reduced dynamic range at higher baseline stiffness, rather than a purely force-dependent effect.

PR showed a related but distinct response. Under normal perfusion, PR decreased progressively with increasing voluntary force. In the BFR/post-release state, this downward force dependence was preserved but shifted from a lower hemodynamic baseline, as reflected by the 41% attenuation of the PR-force slope. Pairwise BFR-related PR reductions were significant at rest and at 15% MVC post-release, but not at 30% MVC, consistent with both the reduced slope magnitude and the elapsed time since cuff release. Thus, PR appears especially sensitive to passive obstruction and early reperfusion, while its incremental contraction-dependent change becomes smaller at higher force. Reactive hyperemia and metabolite-related changes after cuff release may have contributed to the early PR decrease observed here [11, 12, 28]. The non-parallel behavior of SWS and PR supports the view that these parameters provide complementary rather than redundant information about the mechanical state of the tissue.

Men exhibited a larger resting BFR-induced increase in SWS than women, and the reduction in the SWS-force slope was apparent in men but not in women. These patterns may reflect sex- or size-related differences in the acute tissue response to BFR, possibly influenced by limb size, cuff-to-limb geometry, muscle volume, or training status [29, 30]. However, the subgroup sample was modest, and muscle size, habitual training, and local perfusion were not assessed. The apparent sex-related differences therefore remain exploratory and should be tested in larger, sex-balanced cohorts.

The ROC results should be interpreted as an internal characterization of state separation within this experimental dataset. Discrimination was strongest at rest and 15% MVC, where the BFR-related effects were largest, and weak at 30% MVC, where the paired contrasts were smallest. These analyses do not establish diagnostic thresholds and should not be generalized beyond the present protocol.

Several limitations also define the most useful next steps. First, contraction measurements after the hemodynamic perturbation were obtained after cuff release rather than during sustained occlusion. The current data therefore support conclusions about early post-BFR reperfusion states rather than steady-state contraction under active cuff inflation. Second, the fixed condition sequence means that potential fatigue effects cannot be fully excluded, which is relevant because repeated submaximal contractions can themselves alter active muscle stiffness [31]. Third, no electromyographic recordings were obtained, so the degree to which voluntary motor drive was matched across pre-BFR and post-release conditions at the same nominal force target cannot be confirmed. BFR can alter neuromuscular behavior, including motor-unit firing rates and recruitment thresholds during low-load exercise [32], and the absence of EMG prevents ruling out a contribution of altered motor-unit activity to the observed post-release stiffness changes. Fourth, THE cannot disentangle the overlapping effects of increased intraluminal blood pressure, hyperelastic stretching of vessel walls, interstitial fluid shifts, cellular swelling, and metabolite accumulation, all of which may synergistically contribute to changes in SWS and PR. The measured parameters should therefore be interpreted as tissue-level mechanical observables rather than as direct measurements of individual microscopic mechanisms. Fifth, because SWS is derived primarily from wave phase velocity and PR from wave amplitude decay, the two parameters may differ in inversion accuracy and sensitivity to wave attenuation, boundary reflections, and signal-to-noise conditions. Finally, measurements were limited to one muscle. Future studies should combine THE with EMG, direct hemodynamic or metabolic readouts, and fluid-sensitive imaging, acquire time-resolved measurements through the full occlusion and release cycle, and assess multi-muscle responses in larger cohorts.

In summary, multi-parametric THE captured both contractile loading and BFR-related hemodynamic perturbation in the VL. SWS and PR responded differently across force levels and perfusion states, indicating that elastic and dissipative wave metrics provide complementary information about muscle biomechanics. These findings support the view of skeletal muscle as an active, perfused soft tissue whose mechanical state is shaped by the interaction of contractile structures and fluid-containing compartments. Because ultrasound THE is cost-effective, does not require specialized high-frame-rate hardware, and provides spatially resolved full-field-of-view estimates, it may be useful for the non-invasive characterization of living soft tissues under controlled physiological perturbations, with potential applications in exercise physiology and clinical muscle assessment.

## Data and code availability

The datasets generated and analyzed during this study are not publicly available due to ethical restrictions but are available from the corresponding author on reasonable request. Custom THE processing code is available from the corresponding author on reasonable request.

